# Primary care consultation length by deprivation and multimorbidity in England

**DOI:** 10.1101/2020.01.27.20018960

**Authors:** Anya Gopfert, Sarah Deeny, Rebecca Fisher, Mai Stafford

## Abstract

**Background:** Longer GP consultations are recommended as one way of improving care for people with multimorbidity. In Scotland, multimorbid patients in deprived areas do not have had longer consultations though their counterparts in the least deprived areas do. This example of the inverse care law has not been examined in England.

**Aim:** To assess GP consultation length by socioeconomic deprivation and multimorbidity.

**Design and Setting:** Random sample of 1.2 million consultations from 1^st^ April 2014-31^st^ March 2016 for 185,755 adults in England drawn from the Clinical Practice Research Datalink.

**Method:** Consultation duration was derived from time of opening and closing the patient’s electronic record. Mean duration was estimated by multimorbidity level and type, adjusted for number of consultations and other patient and staff characteristics and patient and practice random effects.

**Results:** Consultations lasted 10.9 minutes and mean duration increased with number of conditions. Patients with 6+ conditions had 0.9 (95% CI 0.8, 1.0) minutes longer than those with none. Patients with both mental and physical health condition had 0.5 (95% CI 0.4, 0.5) minutes longer than non-multimorbid patients. However, consultations were 0.5 (95% CI 0.4, 0.5) minutes shorter in the most compared with the least deprived fifth of areas at all levels of multimorbidity.

**Conclusion:** GPs in England spend longer with patients who have more conditions but at all multimorbidity levels, those in deprived areas have less time per GP consultation. Research is needed to assess the impact of consultation length on patient and system outcomes for people with multimorbidity.

## Introduction

Multimorbidity is defined as the co-existence of two or more conditions within an individual. Prevalence estimates depend on the conditions counted but recent studies suggest around 23-27% in the general population (1, 2) -- an estimated 14.2 million people in England (3) – are affected and prevalence is increasing across the UK (4). The risk of multimorbidity increases with advancing age and is strongly linked to socioeconomic position, occurring more frequently and 10-15 years earlier in the most deprived compared with least deprived areas (2). Living with multimorbidity can be challenging and may result in poor quality of life and difficulties with everyday activities (5, 6). People with multimorbidity often require significant time and interaction with health services. Providing care to these individuals can be challenging due to the complexity of intersecting, health and care requirements (7). In addition, around 30% of multimorbid people have both physical and mental health conditions, rising to over 40% in the most deprived fifth of areas (2). People with comorbid physical and mental conditions have more complex care needs and can find it more difficult to manage their conditions(8).

Compared with people who are not multimorbid, people with multimorbidity require more input from the healthcare system. They require a higher number of GP consultations and have an increased likelihood of an emergency admission to hospital (1, 9). There is however, some evidence that if a person is more able to manage their multiple health conditions independently, they have fewer emergency admissions (9, 10). One study in an area of high deprivation showed that more time for complex consultations is associated with increased patient enablement, i.e. ability to self-manage conditions (11). The Royal College of GPs, based on this premise, recommend longer consultations for patients with multimorbidity in order to reduce workload on the broader NHS (12). People living with multimorbidity, likewise, have identified longer primary care appointments as an optimal way of improving the quality of their care(13).

Despite these recommendations, research in Scotland has shown that the greater need of patients with multimorbidity living in the most deprived quarter of areas is not reflected in longer consultation length. This contrasts with the least deprived quarter of areas where those with multimorbidity received longer consultations than those without(14). This is an example of the inverse care law, where the availability of good medical care tends to vary inversely with need and can result in unmet need for health care. Research in Scotland has also demonstrated that although consultation rates increase with deprivation, the social gradients in multimorbidity are much steeper, indicating potentially unmet need. We are not aware of any research examining whether this particular example of the inverse care law also applies in England, though consultation length has been found to be shorter in more deprived areas (15, 16).

We studied the association between GP consultation length and presence of multimorbidity or socioeconomic deprivation in England. We tested whether the difference in consultation length for patients with and without multimorbidity varied between more and less deprived areas in England. We also assessed whether these factors were affected by multimorbidity type.

## Methods

Data were obtained from the Clinical Practice Research Datalink (CPRD), a research database of anonymised patient records covering approximately 6.9% of the UK population (17). Our dataset consisted of a random sample of n=300,000 people in England eligible for linkage to an area-based measure of socioeconomic deprivation and registered between 1^st^ April 2014 and 31^st^ March 2016 (or who died during this period) in an Up-To-Standard practice (i.e. a quality indicator based on continuous recording of patient data and completeness of recorded deaths). We included consultations over this two-year follow-up period. For this study, we excluded those aged under 18 years.

### Consultation duration

Consultation duration was captured in whole minutes and derived from the opening and closing time for a patient’s electronic patient record. We analysed only face-to-face consultations with a GP or GP registrar. We excluded consultations where the record was opened for administrative purposes, telephone consultations (due to the large number which may be triage appointments followed by face-to-face consultations), and home visit consultations (as the recorded duration would only represent the time taken to record the consultation after it has ended). Consultations recorded as lasting over 60 minutes were truncated at 60 mins as these were considered unlikely to reflect actual consultation length (18). Consultations recorded as lasting 0 minutes were set to 0.5 minutes (15).

For the main analysis, we excluded consultations less than two minutes duration as it was deemed these may not reflect accurate consultation length. In sensitivity analysis, we included all consultations irrespective of duration.

### Multimorbidity status

We derived the presence or absence of 36 conditions at the start of follow-up on April 1^st^ 2014. These 36 conditions were identified in previous work because they are likely to be chronic, related to reduced quality of life and mortality risk, and with substantial need for ongoing treatment (1) and used publicly available lists(19) for Read codes (i.e. codes used by UK primary care practitioners to record information about diagnoses) and product codes (i.e. codes specific to CPRD to record information about pharmacological and non-pharmacological products). Patients were grouped according to the number of conditions. In addition, we grouped patients into: those with no or one condition; those with two or more conditions including at least one mental health condition (depression or anxiety, anorexia or bulimia, alcohol problems, other psychoactive substance use, schizophrenia) which we refer to as “multimorbid – including a mental health condition”; and those with two or more physical health conditions, which we refer to as “multimorbid – physical only”.

### Socioeconomic deprivation

Deprivation was based on the patient’s area of residence (Lower Super Output Area level) using deciles of the 2015 Index of Multiple Deprivation(20), grouped into high deprivation (deciles 1-3), medium deprivation (4-7) or low deprivation (8-10). Linkage was undertaken by CPRD.

### Covariates

We included patient and staff factors that can influence consultation duration (15) and may confound an association between duration and deprivation or multimorbidity. Previous work shows women and older people tend to have longer consultations, though the association between duration and age is not linear. More consultations may be used to extend the total consultation time where practice policies only allow for fixed, shorter appointments so we adjusted for number of consultations the patient had during the two-year follow-up. GP registrars are GPs in training and are typically allocated longer duration for their consultations. GP registrars may also not be assigned the most complex patients. Consultations with female health care staff tend to be longer (15) as do consultations in urban areas(20).

### Statistical analysis

We conducted multilevel linear regression analysis with consultation length as the dependent variable and controlled for sex and age, number of GP consultations in the two-year follow-up period, GP trainee status, GP gender, urban-rural classification, index of multiple deprivation and multimorbidity level or type. Three-level regression models accounted for the non-independence of multiple consultations within patients, and patients within practices. We additionally tested for an interaction between index of multiple deprivation and multimorbidity.

Consultation length is not normally distributed but previous studies (15) have analysed it using means and multilevel linear regression models. In sensitivity analysis, we repeated the regression models using multilevel Poisson regression. The direction and statistical significance of the associations of interest were unchanged (results available on request). We therefore present the linear regression results here.

## Results

The original sample of patients aged 18 and over contained data on 2,553,413 face-to-face consultations, of which 1,522,128 were with a GP or GP registrar. Of these, 263,309 lasted less than two minutes. We conducted the main analysis based on 1,258,919 consultations lasting two or more minutes in 190,036 patients. Consultations of less than two-minute duration were more common in those with more conditions (20% of those with 6+ conditions and 13% in those with no conditions; Supplementary Table 1).

Fifty-five per cent of the sample were women, 25.9% lived in the least deprived fifth of areas in England, and 35.5% had two or more conditions (Table 1). Twenty-three per cent had two or more physical conditions and 12.4% had multimorbidity that included at least one mental health condition.

**Table 1.** Characteristics of included patients (n=190,036)

|  | % (N) |
| --- | --- |
| Sex |  |
| Men | 45.0 (86,106) |
| Women | 55.0 (103,930) |
| Age |  |
| 18-29y | 14.5 (27,551) |
| 30-39y | 14.7 (27,913) |
| 40-49y | 18.8 (35,699) |
| 50-59y | 18.2 (34,483) |
| 60-69y | 16.0 (30,399) |
| 70-79y | 11.3 (21,383) |
| 80+y | 6.7 (12,608) |
| Index of Multiple deprivation |  |
| Quintile 1 (least deprived) | 25.9 (49,183) |
| Q2 | 21.1 (40,057) |
| Q3 | 20.3 (38,582) |
| Q4 | 18.4 (34,900) |
| Q5 (most deprived) | 14.4 (27,314) |
| Multimorbidity level |  |
| 0 conditions | 39.2 (74,548) |
| 1 condition | 25.3 (48,043) |
| 2 conditions | 14.9 (28,306) |
| 3 conditions | 9.0 (17,054) |
| 4-5 conditions | 8.4 (15,969) |
| 6+ conditions | 3.2 (6,116) |
| Multimorbidity type |  |
| Not multimorbid<sup>a</sup> | 64.5 (122,591) |
| Multimorbid – physical only<sup>b</sup> | 23.1 (43,822) |
| Multimorbid – including a mental health condition<sup>c</sup> | 12.4 (23,623) |

<sup>a</sup>0-1 long-term condition; <sup>b</sup>2+ physical conditions and no mental health conditions; <sup>c</sup>2+ conditions with at least one mental health condition

In unadjusted analysis (Table 2), women had longer consultations (11.0 minutes) and more consultations (8.6 over 2 years) than men (10.9 minutes and 6.7 consultations respectively). Older people did not have longer consultations, but they had more consultations compared with younger people. Compared with fully qualified GPs, GP registrars had longer consultations with a mean duration of 14.4 minutes. Mean consultation length was shorter for people living in the most compared with the least deprived fifth of areas (10.7 vs 11.2 minutes). Shorter consultations were also seen for patients that were not multimorbid (10.8 minutes compared with 11.0 for multimorbid patients). Among multimorbid patients, those with at least one mental health condition had mean consultation time of 11.1 minutes and those with only physical health conditions 10.9 minutes.

**Table 2.**
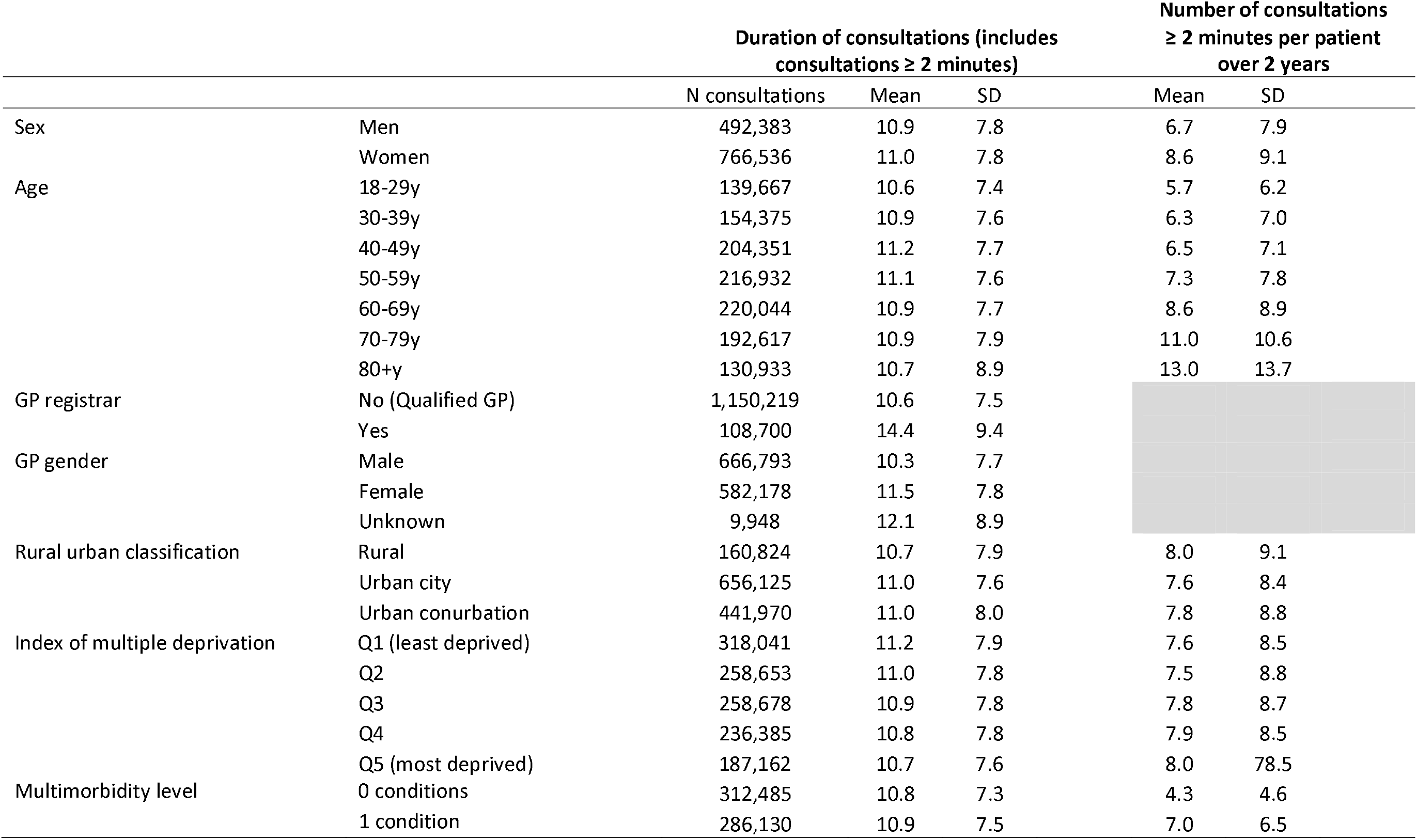
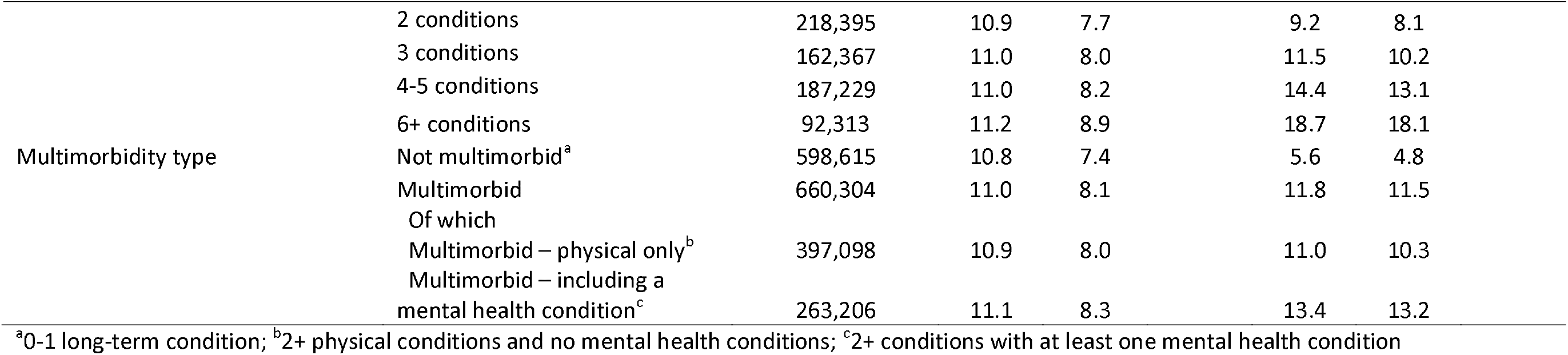
Consultations by sociodemographic characteristics.

Table 3 summarises estimates from the regression models. In the main analysis limited to consultations lasting two minutes or longer, controlling for patient and staff characteristics, residence in a more deprived area was associated with a shorter consultation. Mean duration was 0.46 (95% CI 0.40, 0.53) minutes shorter for those in the most compared with the least deprived fifth of areas. Consultation length increased with number of conditions the patient had and was 0.94 (95% CI 0.84,1.03) minutes longer for those with six or more compared with no long-term conditions. Consultation length also depended on multimorbidity type, with patients with two or more physical conditions having 0.30 minutes longer with the GP and those with two or more conditions including a mental health condition having 0.47 minutes longer compared with non-multimorbid patients (Table 4).

**Table 3.**
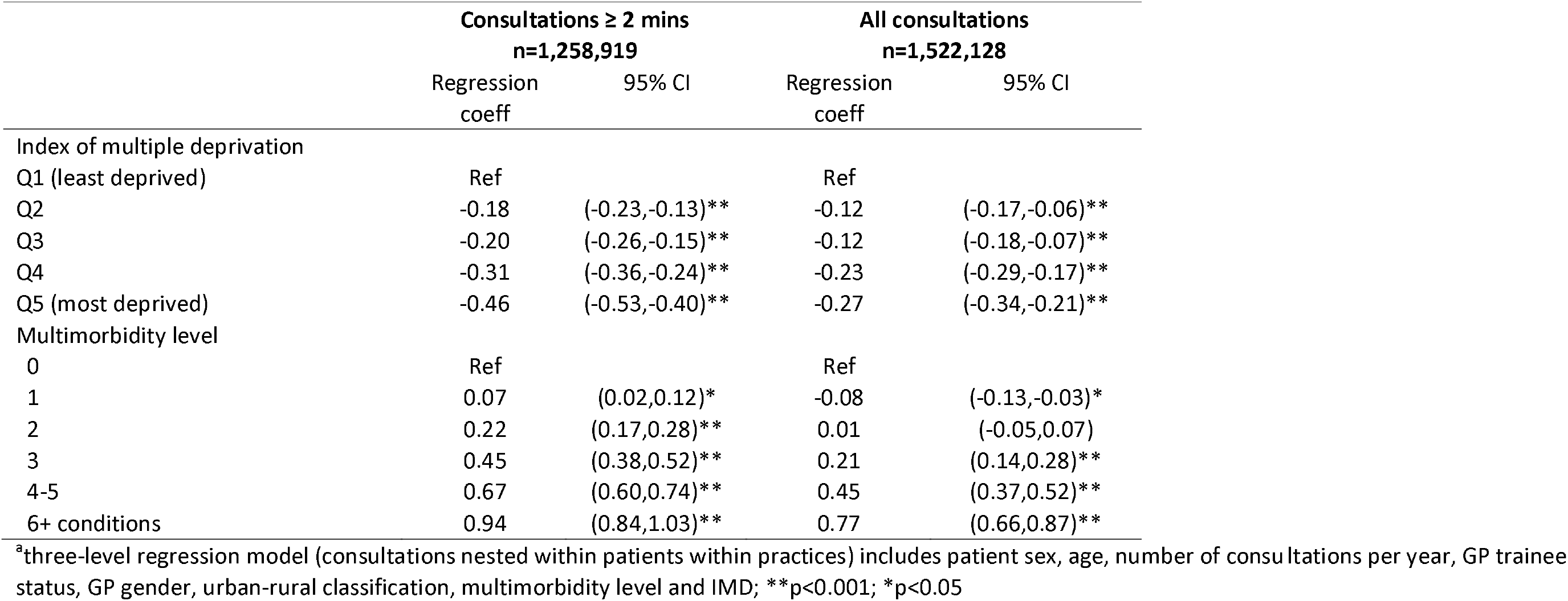
Association^a^ between consultation duration and multimorbidity level and area deprivation.

|  | Consultations ≥ 2 mins<br>n=1,258,919 |  | All consultations<br>n=1,522,128 |  |
| --- | --- | --- | --- | --- |
|  | Regression<br>coeff | 95% CI | Regression<br>coeff | 95% CI |
| Index of multiple deprivation |  |  |  |  |
| Q1 (least deprived) | Ref |  | Ref |  |
| Q2 | -0.18 | (-0.23,-0.13)** | -0.12 | (-0.17,-0.06)** |
| Q3 | -0.20 | (-0.26,-0.15)** | -0.12 | (-0.18,-0.07)** |
| Q4 | -0.31 | (-0.36,-0.24)** | -0.23 | (-0.29,-0.17)** |
| Q5 (most deprived) | -0.46 | (-0.53,-0.40)** | -0.27 | (-0.34,-0.21)** |
| Multimorbidity level |  |  |  |  |
| 0 | Ref |  | Ref |  |
| 1 | 0.07 | (0.02,0.12)* | -0.08 | (-0.13,-0.03)* |
| 2 | 0.22 | (0.17,0.28)** | 0.01 | (-0.05,0.07) |
| 3 | 0.45 | (0.38,0.52)** | 0.21 | (0.14,0.28)** |
| 4-5 | 0.67 | (0.60,0.74)** | 0.45 | (0.37,0.52)** |
| 6+ conditions | 0.94 | (0.84,1.03)** | 0.77 | (0.66,0.87)** |

<sup>a</sup>three-level regression model (consultations nested within patients within practices) includes patient sex, age, number of consultations per year, GP trainee status, GP gender, urban-rural classification, multimorbidity level and IMD; \*\*p<0.001; \*p<0.05

**Table 4.** Association^a^ between consultation duration and multimorbidity type and area deprivation.

|  | Consultations ≥ 2 minutes<br>n=1,258,919 |  | All consultations<br>n=1,522,128 |  |
| --- | --- | --- | --- | --- |
|  | Regression<br>coeff | 95% CI | Regression<br>coeff | 95% CI |
| Index of multiple deprivation |  |  |  |  |
| Q1 (least deprived) | Ref |  | Ref |  |
| Q2 | -0.18 | (-0.23,-0.12)** | -0.11 | (-0.17,-0.06) |
| Q3 | -0.19 | (-0.25,-0.14)** | -0.11 | (-0.17,-0.06)* |
| Q4 | -0.29 | (-0.35,-0.23)** | -0.22 | (-0.27,-0.16)** |
| Q5 (most deprived) | -0.45 | (-0.51,-0.38)** | -0.26 | (-0.32,-0.19) |
| Multimorbidity type |  |  |  |  |
| Not multimorbid<sup>b</sup> | Ref |  | Ref |  |
| Multimorbid – physical only<sup>c</sup> | 0.30 | (0.25,0.35)** | 0.19 | (0.14,0.24)** |
| Multimorbid – including a mental<br>health condition<sup>d</sup> | 0.47 | (0.42,0.53)** | 0.29 | (0.24,0.35)** |

<sup>a</sup>three-level regression model (consultations nested within patients within practices) includes patient gender, age, number of consultations per year, GP trainee status, GP gender, urban-rural classification, multimorbidity type and IMD; <sup>b</sup>0-1 long-term condition; <sup>c</sup>2+ physical conditions and no mental health conditions; <sup>d</sup>2+ conditions with at least one mental health condition; \*\*p<0.001; \*p<0.05

We found no clear evidence that the association between multimorbidity level or type and consultation length was different for patients in more versus less deprived areas. Figure 1 shows consultation duration by index of multiple deprivation and multimorbidity type from the model including the interaction of these two factors. It illustrates that patients in the most deprived areas had shorter consultations than those in the least deprived areas for all multimorbidity types. It also illustrates that the mean consultation length for a non-multimorbid patient in a low deprivation area (10.9 minutes) was the same as that for a multimorbid patient with physical and mental health conditions in an area of high deprivation.

**Figure 1.**
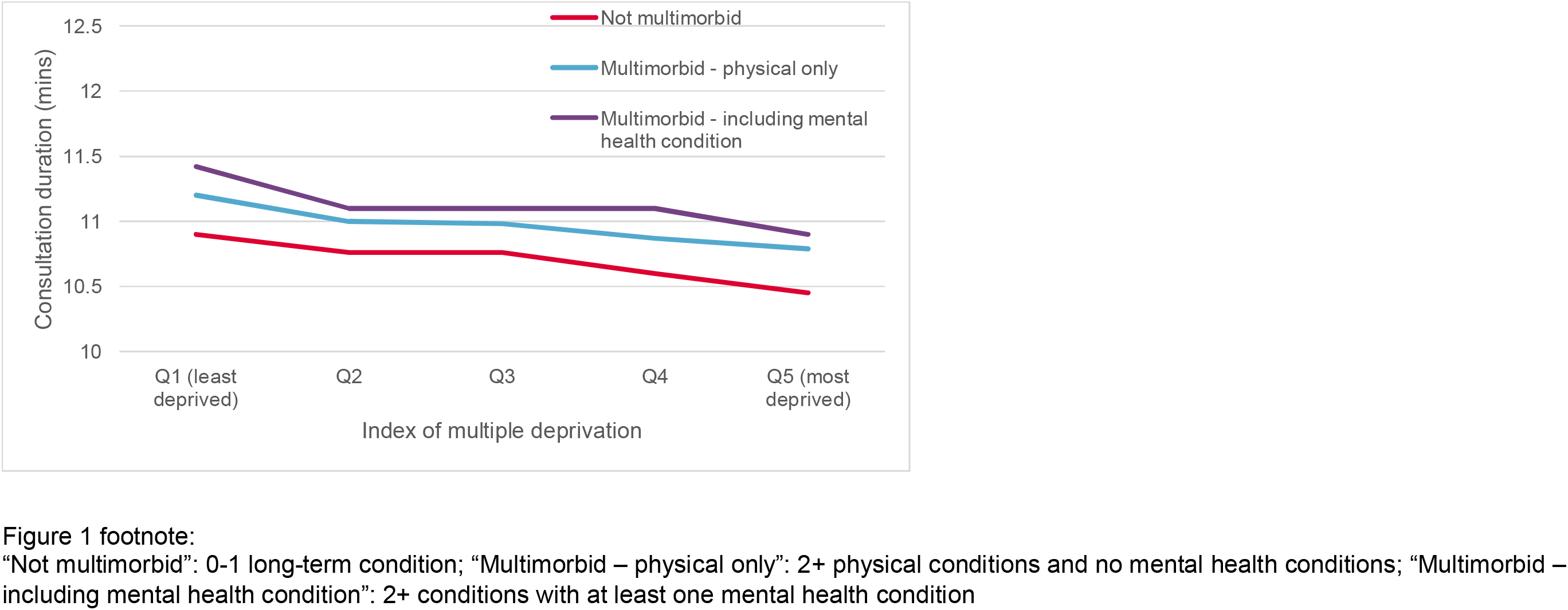
Consultation duration by index of deprivation and multimorbidity type: consultations lasting two minutes or more.

The same patterns were found when all consultations (including those lasting less than two minutes) were analysed. Regression estimates show smaller differences in consultation length when all consultations were included, as expected because very short consultations were more common in patients with more long-term conditions.

## Discussion

### Summary

Living in an area of high socioeconomic deprivation is associated with shorter GP consultations. GP consultation length increased with increasing number of health conditions. Consultations were also longer for multimorbid patients with a mental health condition than for multimorbid patients with physical conditions only.

### Strengths and limitations

A strength of this study was the large sample size and the use of routine data to minimise selection bias. The use of multilevel regression analysis allowed for unobserved similarities between practices and between patients that could affect consultation duration. The association between deprivation and duration remained on adjustment for total number of consultations, indicating that use of additional consultations did not explain the shorter consultations in more deprived areas. This study was limited by several factors. CPRD data provides consultation time based on the open and close time of the electronic record. This is the amount of time a practitioner had the file open, which may be affected by other factors including practitioner preference regarding whether to complete and close a record while the patient is present or later in the day and the possibility of clinicians forgetting to close a consultation until after the care episode has ended (though we capped all consultations at a maximum of 60 minutes). However, there is no evidence to suggest that these factors differ by patient level of deprivation or multimorbidity. Previous analysis used video-recording to accurately capture consultation duration though this approach may have altered GP behaviour. This study also focuses on primary care delivered by GPs. Future analysis should also consider consultations with nurses, since nurses provide a sizeable proportion of primary care for people with multimorbidity(18). As primary care only forms a single component of health care for people with multimorbidity, further study should repeat this analysis within secondary care and other parts of the health system.

The adjusted difference in consultation time for patients in the most compared with the least deprived areas amounted to 0.5 minutes. The magnitude of this difference appears small and further work is needed to assess whether consultation length is associated with poor experience, poor outcomes or greater use of other health services, as others have noted(21). This small value should however be interpreted in the context of an average consultation of just under 11 minutes.

### Comparison with existing literature

This paper supports evidence that multimorbidity and deprivation influence consultation time with a GP. Particularly concerning is ongoing evidence indicating that patients in deprived areas have shorter consultation times (15). Our findings confirm evidence previously found in Scotland based on videotaped consultations to provide an accurate measure of time spent with patients(14). That study considered a single consultation for each patient and our study adds to their findings in showing that a similar pattern is observed (i.e. shorter consultations for patients in more deprived areas) across multiple consultations over a two-year period of usual care. This is likely to reflect ongoing job pressures for GPs in deprived areas, and a greater need for care among this group of patients(. Practices in deprived areas tend to have lower levels of GP staffing(22). The staffing level and patient load at a particular GP surgery influences the work pressure for GPs, and therefore can influence the consultation length time available(22). Given these pressures, other factors that could affect patient experience and patient outcomes, such as continuity of care or GP empathy, might also differ by deprivation level, as has been found in Scotland (15). We did not examine these other characteristics of the consultation, and further work is needed to explore those factors and to test the contribution of consultation length, continuity of care and patient experiences of the consultation to outcomes. The previous study set in Scotland compared consultation length for practices in high and low deprivation areas whereas our study used deprivation in the patient’s local area. Although patient and practice deprivation will be positively correlated, they may influence consultation length independently via different mechanisms. Analysis including deprivation at both patient and practice level would be useful to explore this further but was not possible with the current data.

We also identify that patients with multimorbidity receive longer consultations. Consultation length increased with the number of conditions a patient had. This is in line with calls for longer GP consultations for multimorbid patients, though whether these relatively small differences in consultation length are related to, or sufficient to achieve better patient outcomes remains to be tested. Previous evidence from Scotland (14) showed that patients with multimorbidity received around three minutes longer with their GP than those without multimorbidity in affluent areas but this was not the case in deprived areas. In England, multimorbid patients in more and less deprived areas had longer consultations than their non-multimorbid counterparts.

GPs in England are spending longer with patients who have more long-term conditions. The consultation length also depends on the types of conditions the patient has. Having a mental health condition can make it more difficult to manage complex care needs, and longer consultations have been linked to better handling of psychological problems in primary care (23, 24). Our analysis shows that multimorbidity including a mental health condition was associated with having a longer consultation compared with having multiple physical conditions and compared with not having multimorbidity. However, the analysis also shows that this additional time is counteracted by living in a deprived area. The association between deprivation and consultation length is equal in magnitude and opposite in effect to the association between multimorbidity and consultation length. This means that a patient with multiple mental and physical health conditions living in an area of high deprivation receives the same amount of time with their GP as a non-multimorbid person in an area of low deprivation.

### Implications

Our study shows that the inverse care law is alive and well in general practice in England. Not only do people living in more deprived areas of the country have on average shorter GP appointments – the same pattern is observed even when those people have multiple health conditions. The findings suggest that there could be unmet need among patients with complex care needs, particularly patients with both mental and physical health conditions, living in deprived areas.

Understanding *why we* observe shorter consultation times in general practice in areas of high deprivation is crucial to understanding *how* this could be changed. This includes understanding patient factors as well as those related to the organisation and delivery of general practice.

Undersupply of GPs relative to population need, and corresponding higher workload may be a key driver of shorter consultation times, and evaluation of the impact of initiatives encouraging GPs to train and work in under-doctored areas is awaited. Increasing skill mix in primary care by recruiting additional allied health professionals is seen as one way of freeing up GP time to focus on more complex patients. These staff may also directly contribute to and improve care for people with multimorbidity. Initiatives to ensure these additional staff will be distributed equitably across the country and to encourage them to work in areas of high deprivation will be needed. If additional staff gravitate to areas of lower deprivation, then there will be paradoxically even fewer staff relative to need in the areas of highest deprivation (22).

The positive association between consultation length and number of long-term conditions that we identified is in line with calls for longer GP consultations for multimorbid patients, though whether these relatively small differences in consultation length are related to, or sufficient to achieve better patient outcomes remains to be tested.

### Conclusion

GPs in England are spending longer with patients who have more long-term conditions. They are also spending longer with patients who have complex care needs including mental as well as physical health conditions. However, consultation length is shorter in more deprived areas. This is the case for multimorbid as well as non-multimorbid patients. This fully counteracts the additional time given to multimorbid patients meaning that patients with multiple mental and physical health conditions living in the most deprived fifth of areas receive no longer with their GP than a non-multimorbid person in an area of low deprivation.

Continued monitoring of the distribution of the primary care workforce by socioeconomic deprivation and how this relates to consultation length for more complex patients will be needed. Further research is also needed to assess the impact of consultation length on patient and system outcomes for people with multimorbidity. Evidence from this study should bolster calls on policy makers to ensure that national policy related to general practice is designed to achieve equity of care, so that care in general practice can be delivered in proportion with need.

## Data Availability

We used data from the Clinical Practice Research Datalink (CPRD). Data access for this project has been approved (ISAC 17_150R). Data used for this analysis is not publically available but anonymised patient datasets can be extracted for researchers against specific study specifications, following protocol approval from the Independent Scientific Advisory Committee (ISAC) https://www.cprd.com/

## Funding

This study was funded by The Health Foundation as part of core activity of members of staff at The Health Foundation.

## Ethical approval

Routinely collected, retrospective, anonymised data were used for this analysis. Approval to use the data was granted by the Independent Scientific Advisory Committee (CPRD protocol number ISAC17_150RMn2).

## Competing interests

Authors have no competing interests.

## Acknowledgements

We thank the Data Management Team at the Health Foundation for their work in preparing the data.

---

**Supplementary Table 1.** Frequency of short consultations by multimorbidity and deprivation.

|  | % consultations lasting <2 mins |
| --- | --- |
| Multimorbidity level |  |
| 0 | 13.3 |
| 1 | 16.4 |
| 2 | 17.9 |
| 3 | 19.0 |
| 4-5 | 19.5 |
| 6+ consultations | 20.2 |
| Multimorbidity type |  |
| Not multimorbid<sup>a</sup> | 14.8 |
| Multimorbid – physical only<sup>b</sup> | 19.1 |
| Multimorbid – including a mental health condition<sup>c</sup> | 18.7 |
| Index of multiple deprivation |  |
| Q1 (least deprived) | 22.3 |
| Q2 | 20.4 |
| Q3 | 20.2 |
| Q4 | 20.4 |
| Q5 (most deprived) | 18.3 |

“Not multimorbid”: 0-1 long-term condition; “Multimorbid – physical only”: 2+ physical conditions and no mental health conditions; “Multimorbid – including mental health condition”: 2+ conditions with at least one mental health condition

## Notes

### Competing Interest Statement

The authors have declared no competing interest.

